# Recording of Pharmacy First consultations in general practice records in England: an observational study of the service’s first year using OpenSAFELY

**DOI:** 10.1101/2025.09.30.25336964

**Authors:** Viveck J Kingsley, Milan Wiedemann, Christopher Wood, Helen J Curtis, Amelia Green, Louis Fisher, Colm D Andrews, David Evans, Amir Mehrkar, Sebastian Bacon, Ben Goldacre, Amelia C Taylor, Diane Ashiru-Oredope, Kimberley Sonnex, Thomas Allen, Hannah Higgins, Tracey Thornley, Nicholas Mays, Rebecca E Glover, Rachel A Elliott, Anthony J Avery, Brian MacKenna

## Abstract

**Background:** Pharmacy First, a national community pharmacy service, launched in January 2024 to improve access to primary care for patients with minor conditions facing backlogs caused by the COVID-19 pandemic. Pharmacies are required to share details about their consultations with general practices. We aimed to describe how and what clinical activity was recorded in general practice during the first year of the service.

**Methods:** With the approval of NHS England, we conducted a retrospective cohort study using OpenSAFELY-TPP including Pharmacy First consultations between 31 January 2024 and 30 January 2025. We described patient demographics, consultation trends, and the clinical conditions and medications coded with Pharmacy First consultations.

**Results:** A total of 402,165 Pharmacy First consultations were recorded for 340,710 patients from a general population of 26,142,380 registered patients in OpenSAFELY-TPP. Acute pharyngitis (28.9%) and uncomplicated urinary tract infection (28%) were the most frequently recorded conditions. By January 2025, 36.3% of recorded Pharmacy First consultations had a clinical condition, medication, or both. Females, younger adults and those living in more deprived areas were observed more often in Pharmacy First records compared to the general population.

**Conclusion:** Increasing recording of the Pharmacy First community pharmacy service was observed in general practice records during its first year, particularly among younger and more deprived populations. However, variation in structured recording of consultation details may limit evaluation.

## Background

The COVID-19 pandemic placed sustained pressure on the National Health Service (NHS) in England, with persistent backlogs and longer waiting times for general practice (GP) appointments (1,2). As a result, community pharmacies have increasingly adopted clinical functions (3). As part of the NHS COVID-19 recovery plan (4), NHS England launched the Pharmacy First service on 31st January 2024, enabling community pharmacists to assess for and manage seven common conditions, and supply prescription-only medications, where appropriate (5,6).

Pharmacy First requires sharing of clinical information between GPs and community pharmacies. NHS England mandated use of *GP Connect: Update Record,* which enables registered pharmacies to send summaries of Pharmacy First consultations directly into GP records (7) using pre-specified Systematized Nomenclature of Medicine Clinical Terms (SNOMED CT) codes (8). This structured transfer aims to ensure consistency, accuracy and speed of recording Pharmacy First consultations in GP records to support GP clinical decision-making when there is escalation back to GPs (9). However, its rollout has been gradual, with community pharmacy and GP software systems being ready at different timepoints (10).

This study forms part of the National Institute for Health and Care Research (NIHR)-commissioned review of the implementation and impact of Pharmacy First (11), using the OpenSAFELY platform for near real-time access to pseudonymised GP records (12,13).

We aimed to describe how Pharmacy First Clinical Pathway activity was recorded in OpenSAFELY-TPP during its first year. We report monthly activity compared to publicly reported figures, assess the information on clinical conditions and medications transferred to GP records, and describe Pharmacy First users’ demographics.

## Methods

### Study design and population

We conducted a retrospective cohort study, using routinely collected electronic health record (EHR) data in OpenSAFELY-TPP between 31st January 2024 and 30th January 2025, the first year of Pharmacy First. We included all individuals who were alive and registered at a TPP practice during the study period, male or female and aged 0-120 years. We did not apply eligibility criteria defined for Pharmacy First pathways as we sought to describe all consultations as they appeared in GP records (5).

### Data sources

#### OpenSAFELY-TPP

All data were stored and analysed securely using the OpenSAFELY platform (https://www.opensafely.org/), as part of the NHS England OpenSAFELY COVID-19 service. Data include pseudonymised data such as coded diagnoses, medications and physiological parameters. No free text data are included. No GP data from patients who have registered a Type-1 Opt out with their GP surgery were included in this study.

#### NHS Business Services Authority (BSA) data

For comparison we used the publicly available Pharmacy and appliance contractor dispensing dataset which captures all monthly Pharmacy First consultations reimbursed in England (14).

#### Identification of clinical events and medications

We extracted Pharmacy First consultations together with associated clinical conditions and medications (Box 1).

##### Box 1.

Code written to identify clinical conditions and medications of Pharmacy First consultations in OpenSAFELY. Using Electronic Health Records Query Language (ehrQL) (https://docs.opensafely.org/ehrql/) we identified all Pharmacy first consultations through consultation identifiers then extracted all clinical event codes (i.e. conditions) and medications associated with the consultation.

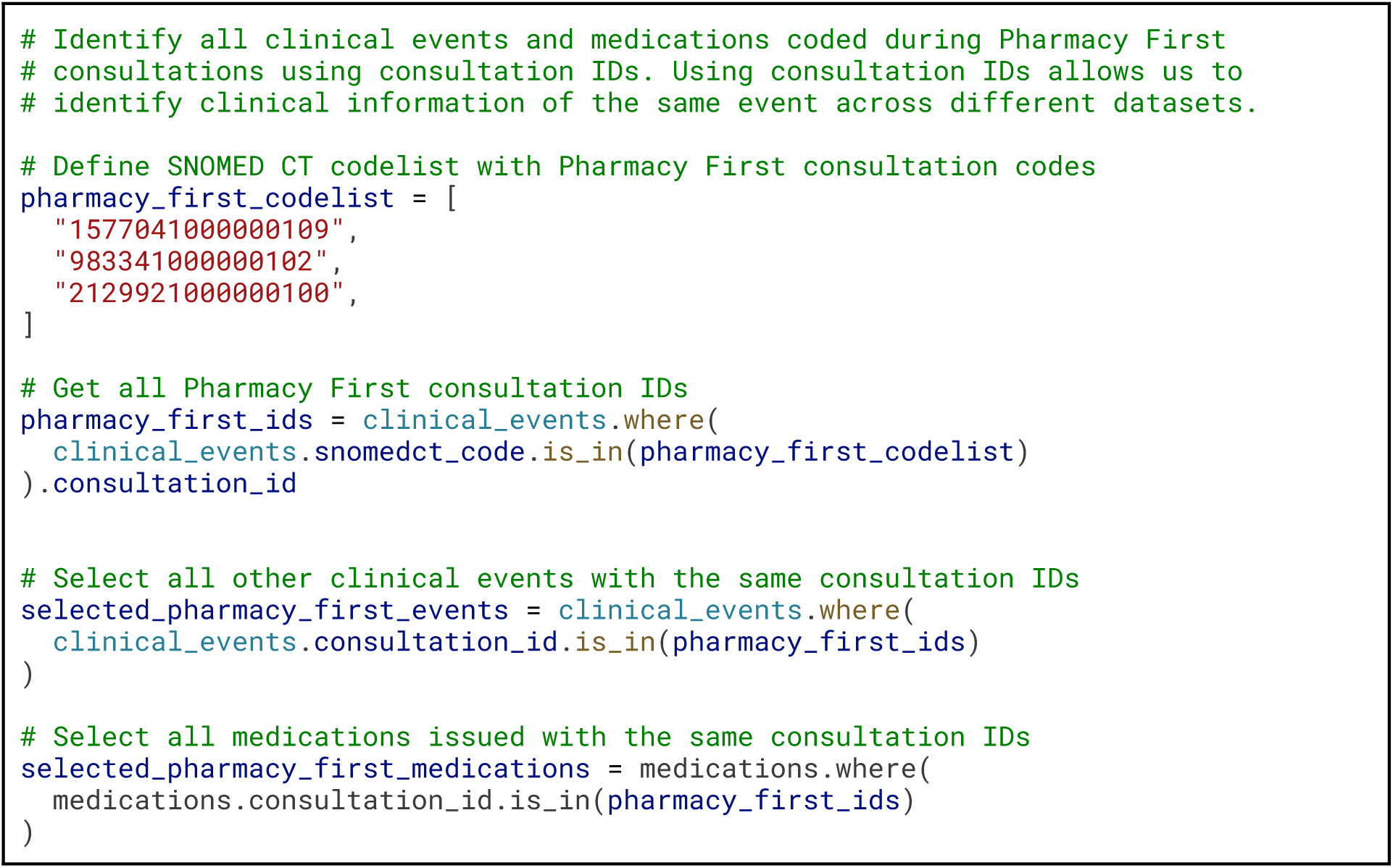

#### Pharmacy First consultations

We used SNOMED CT codes specified by NHS England in the *GP Connect: Update Record* documentation: *Community Pharmacist Consultation Service for minor illness* (1577041000000109) (8). Using *OpenCodelists* (https://www.opencodelists.org/), we identified two further SNOMED CT codes: *Pharmacy First Service* (983341000000102) and *Community Pharmacy Pharmacy First Service* (2129921000000100) (supplementary Table A1).

#### Pharmacy First clinical conditions

Pharmacy First provides clinical pathways for seven conditions: acute otitis media, impetigo, infected insect bites, herpes zoster, acute sinusitis, acute pharyngitis, and uncomplicated urinary tract infection (5). These were identified using SNOMED CT codelists published by NHS England (supplementary Table A1).

#### Pharmacy First medications

We developed medication codelists, using the dictionary of medicines and devices (dm+d) terminology for each clinical condition, to specify the exact drug, formulations and strengths that may be supplied (6) (supplementary Table A1).

### Data analyses

#### Patient population using Pharmacy First

We report demographic (10-year age bands, sex, ethnicity in 16 categories) and regional (Indices of Multiple Deprivation quintiles, region) breakdowns for service users compared to the total registered population. To reduce disclosure risks, counts ≤7 are redacted, then all counts rounded to the nearest 5.

#### Monthly Pharmacy First consultations

Monthly counts of patients with at least one recorded Pharmacy First consultation in GP records are reported, split by Pharmacy First code. We compared monthly Pharmacy First consultation counts in OpenSAFELY-TPP (covering 40% of England’s GP-registered population) with publicly available figures derived from reimbursement claims (100% coverage) (14). We also break down by clinical condition.

#### Pharmacy First consultation information in GP records

Pharmacy First consultations were categorised as: (i) having a recorded Pharmacy First clinical condition; (ii) having a Pharmacy First medication prescribed; or (iii) both; the remainder having neither recorded. Monthly proportions were calculated. We included all three Pharmacy First consultation codes described above, and separately limited to the recommended code, to examine variation in recording practices.

#### Clinical conditions recorded in Pharmacy First consultations

For all Pharmacy First consultations we extracted all SNOMED CT codes recorded which matched the seven NHS England-defined clinical conditions. For each clinical condition, counts were broken down by sex and Index of Multiple Deprivation (IMD). We also compare the percentage of Pharmacy First consultations attributed to each of the seven clinical conditions between OpenSAFELY-TPP and NHS BSA data.

#### Medications recorded in Pharmacy First consultations

Medications issued during Pharmacy First consultations were identified using dm+d codes recorded within the same consultation. We report the ten most frequently recorded generic products (Virtual Medicinal Product [VMP] dm+d level). We describe the percentage of consultations with and without an NHS England-specified medication for each condition.

### Software and reproducibility

Data management was performed using Python v3.9.1 and R v4.0.5. Code for data management and analysis, as well as codelists, are available for review and reuse under MIT open license at https://github.com/opensafely/pharmacy-first/. Detailed pseudonymised patient data is potentially re-identifiable and therefore not shared.

### Patient and public involvement

OpenSAFELY has involved patients and the public in various ways (https://opensafely.org), including two citizen juries exploring public trust; co-developed explainer video (https://www.opensafely.org/about/); patient representation on our OpenSAFELY Oversight Board; partnership with Understanding Patient Data to produce lay explainers; various online public engagement events; and more. The wider Pharmacy First evaluation will involve PPIE and qualitative interviews with patients.

## Results

### Patient population using Pharmacy First

Between 31 January 2024 and 30 January 2025, we identified 402,165 Pharmacy First consultations for 340,710 patients from 26,142,380 registered patients in OpenSAFELY-TPP (Table 1). Pharmacy First users were predominantly female (67.3%) and over half were aged under 40 (59.3%). Most patients were of White British ethnicity (71.3%), and uptake was highest among those in the most deprived quintile (25.4%). Yorkshire and the Humber was the most represented, accounting for 23.7% of Pharmacy First users compared to 14.5% of the registered population.

**Table 1.** Description of patient population with at least one Pharmacy First consultation code (n = 340,710) and total registered population in OpenSAFELY-TPP (N = 26,142,380).

|  | Pharmacy First |  | OpenSAFELY-TPP |  |
| --- | --- | --- | --- | --- |
|  | Count | Percent | Count | Percent |
| **Sex** |  |  |  |  |
| Female | 229,190 | 67.3% | 13,011,745 | 49.8% |
| Male | 111,520 | 32.7% | 13,130,635 | 50.2% |
| **Age** |  |  |  |  |
| 0 - 19 | 94,895 | 27.9% | 5,775,555 | 22.1% |
| 20 - 39 | 106,810 | 31.4% | 7,059,395 | 27.0% |
| 40 - 59 | 86,015 | 25.2% | 6,780,960 | 25.9% |
| 60 - 79 | 45,450 | 13.3% | 5,176,600 | 19.8% |
| 80+ | 7,545 | 2.2% | 1,349,865 | 5.2% |
| **Ethnicity** |  |  |  |  |
| Asian or Asian British |  |  |  |  |
| Bangladeshi | 2,735 | 0.8% | 172,475 | 0.7% |
| Indian | 12,355 | 3.6% | 983,510 | 3.8% |
| Pakistani | 14,720 | 4.3% | 715,395 | 2.7% |
| Any other Asian background | 7,110 | 2.1% | 581,280 | 2.2% |
| Black or Black British |  |  |  |  |
| African | 7,290 | 2.1% | 641,700 | 2.5% |
| Caribbean | 1,635 | 0.5% | 136,735 | 0.5% |
| Any other Black background | 1,650 | 0.5% | 143,570 | 0.5% |
| Mixed |  |  |  |  |
| White and Asian | 1,690 | 0.5% | 121,910 | 0.5% |
| White and Black African | 1,250 | 0.4% | 103,770 | 0.4% |
| White and Black Caribbean | 1,980 | 0.6% | 120,470 | 0.5% |
| Any other mixed background | 2,690 | 0.8% | 216,510 | 0.8% |
| Other ethnic groups |  |  |  |  |
| Chinese | 1,160 | 0.3% | 247,465 | 0.9% |
| Any other ethnic group | 5,485 | 1.6% | 500,815 | 1.9% |
| White |  |  |  |  |
| British | 242,975 | 71.3% | 17,532,065 | 67.1% |
| Irish | 1,475 | 0.4% | 126,620 | 0.5% |
| Any other White background | 26,810 | 7.9% | 2,550,950 | 9.8% |
| (Missing) | 7,695 | 2.3% | 1,247,135 | 4.8% |
| **IMD** |  |  |  |  |
| 1 - Most deprived | 86,690 | 25.4% | 5,183,155 | 19.8% |
| 2 | 67,620 | 19.9% | 5,009,215 | 19.2% |
| 3 | 63,645 | 18.7% | 5,307,215 | 20.3% |
| 4 | 56,835 | 16.7% | 4,936,940 | 18.9% |
| 5 - Least deprived | 48,045 | 14.1% | 4,534,735 | 17.3% |
| (Missing) | 17,875 | 5.2% | 1,171,120 | 4.5% |
| **Region** |  |  |  |  |
| East | 64,230 | 18.9% | 5,929,050 | 22.7% |
| East Midlands | 45,515 | 13.4% | 4,528,605 | 17.3% |
| London | 20,615 | 6.0% | 1,757,575 | 6.7% |
| North East | 23,280 | 6.8% | 1,261,870 | 4.8% |
| North West | 26,785 | 7.9% | 2,402,230 | 9.2% |
| South East | 20,085 | 5.9% | 1,703,325 | 6.5% |
| South West | 42,345 | 12.4% | 3,633,710 | 13.9% |
| West Midlands | 15,850 | 4.7% | 1,042,385 | 4.0% |
| Yorkshire and The Humber | 80,785 | 23.7% | 3,791,585 | 14.5% |
| (Missing) | 1,225 | 0.4% | 92,045 | 0.4% |

Notes. IMD = Index of Multiple Deprivation.

### Monthly Pharmacy First consultations in GP records

Of the 402,165 Pharmacy First consultations, 209,900 (52.2%) were recorded using the SNOMED CT code for *Pharmacy First Service*, and 192,265 (47.8%) using the recommended code, *Community Pharmacist Consultation Service for minor illness* (Figure 1). The code for *Community Pharmacy First Service* was not used. Use of the recommended code increased from around 5,000 per month to 37,565 in December 2024. Use of the *Pharmacy First Service* code remained comparatively stable.

**Figure 1.**
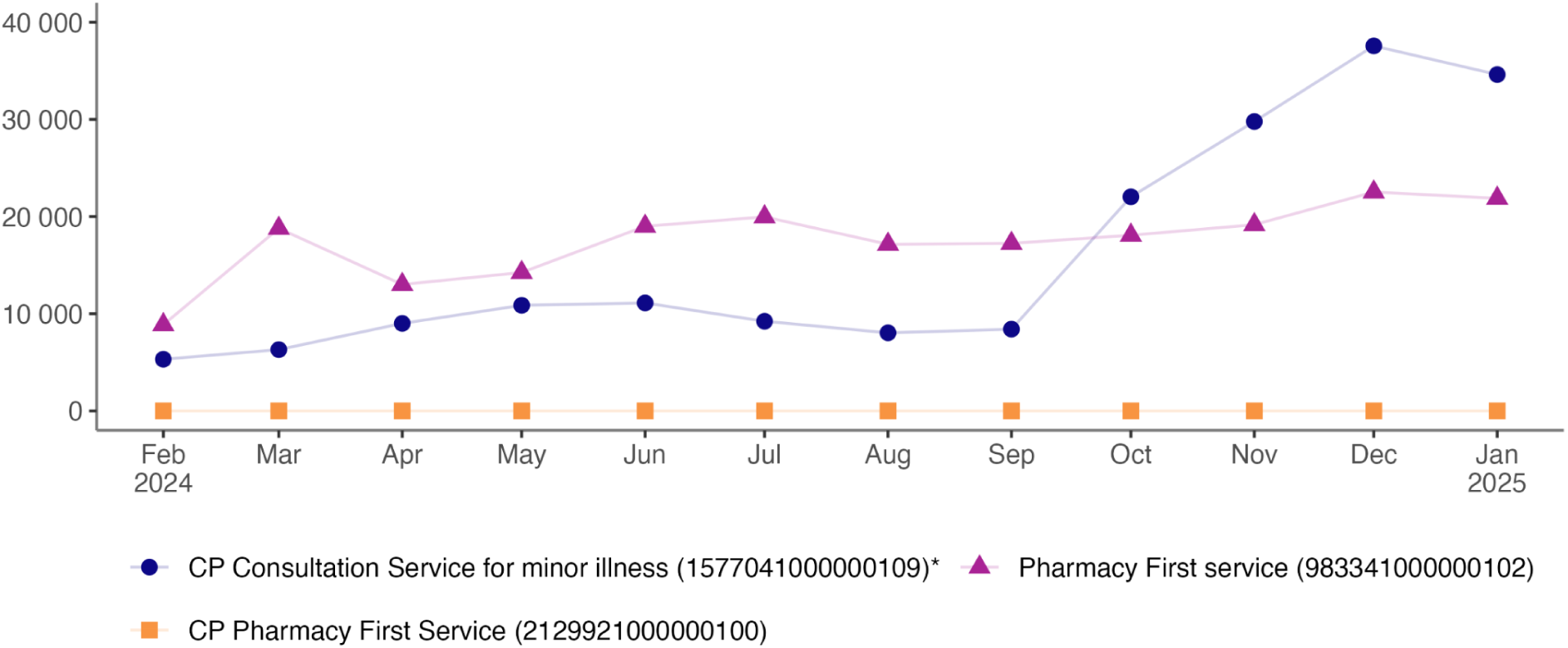
Monthly count of consultations with a Pharmacy First consultation code in their GP records in OpenSAFELY-TPP, broken down by individual Pharmacy First consultation codes. CP = Community Pharmacy. *Recommended Pharmacy First consultation code.

### Comparison with NHS BSA data

The number of Pharmacy First consultations recorded in OpenSAFELY-TPP was substantially lower than expected based on its approximate 40% population coverage. OpenSAFELY-TPP captured between 11.3% and 23.7% of the total consultations reported per month in NHS BSA data (Figure 2). Both datasets showed broadly similar overall trends.

**Figure 2.**
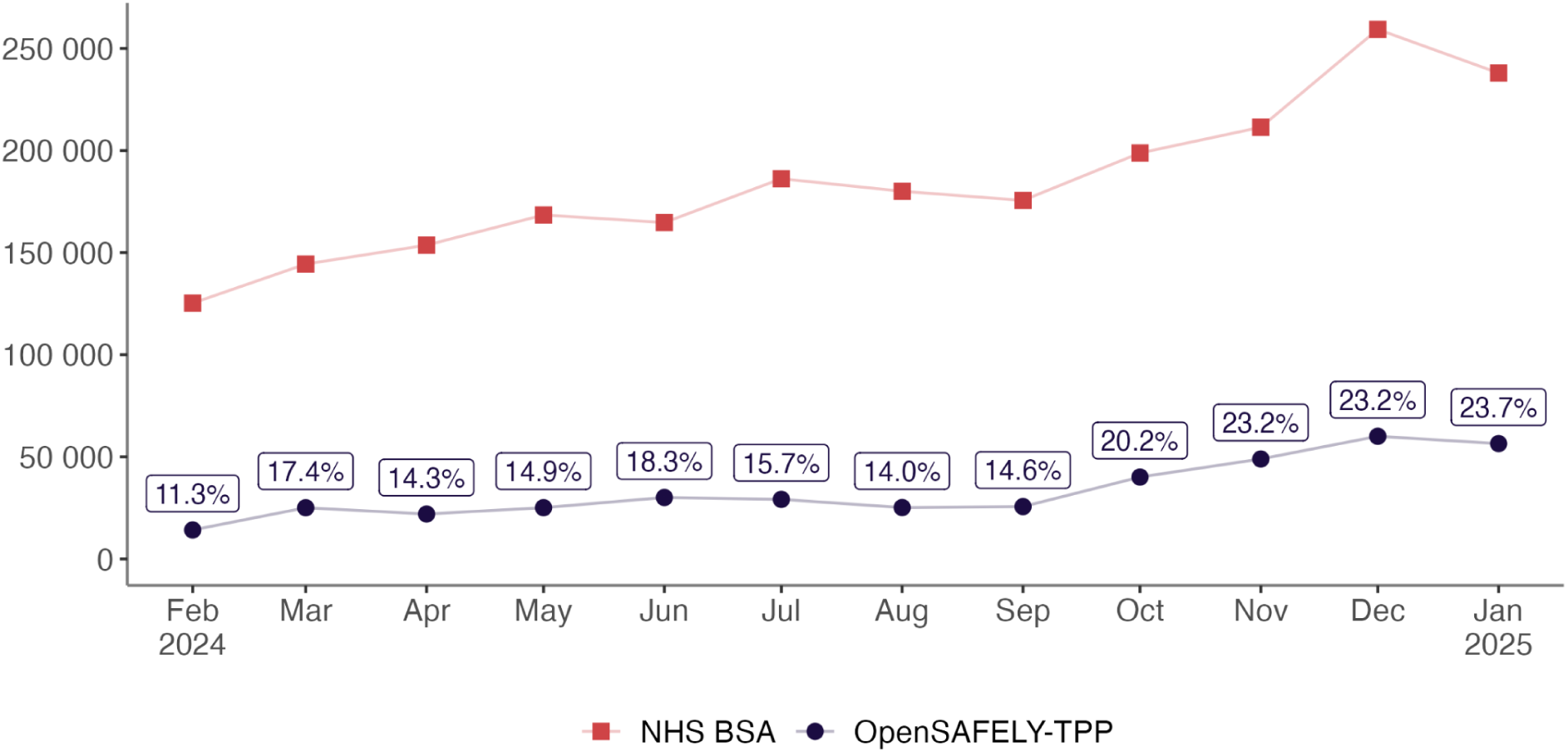
Monthly counts of Pharmacy First consultations in OpenSAFELY-TPP, which represents approximately 40% of the population, and NHS BSA data which represents 100% of reimbursement claims. Labels indicate the percentage of OpenSAFELY-TPP consultations of the BSA total.

### Pharmacy First consultation information

In February 2024, only 4.5% of Pharmacy First consultations in OpenSAFELY-TPP contained a clinical condition code and 0.7% a medication, with both recorded in <0.1% (Figure 3a). By January 2025, 13.5% included a condition, 0.8% a medication, and 22.0% both - totalling 36.3% of consultations with a recorded condition and/or medication. When restricted to consultations recorded using the recommended code (Figure 3b), the overall pattern was similar, but by January 2025, 53.7% had a recorded condition and/or medication (17.8% condition, 0.2% medication, 35.7% both).

**Figure 3.**
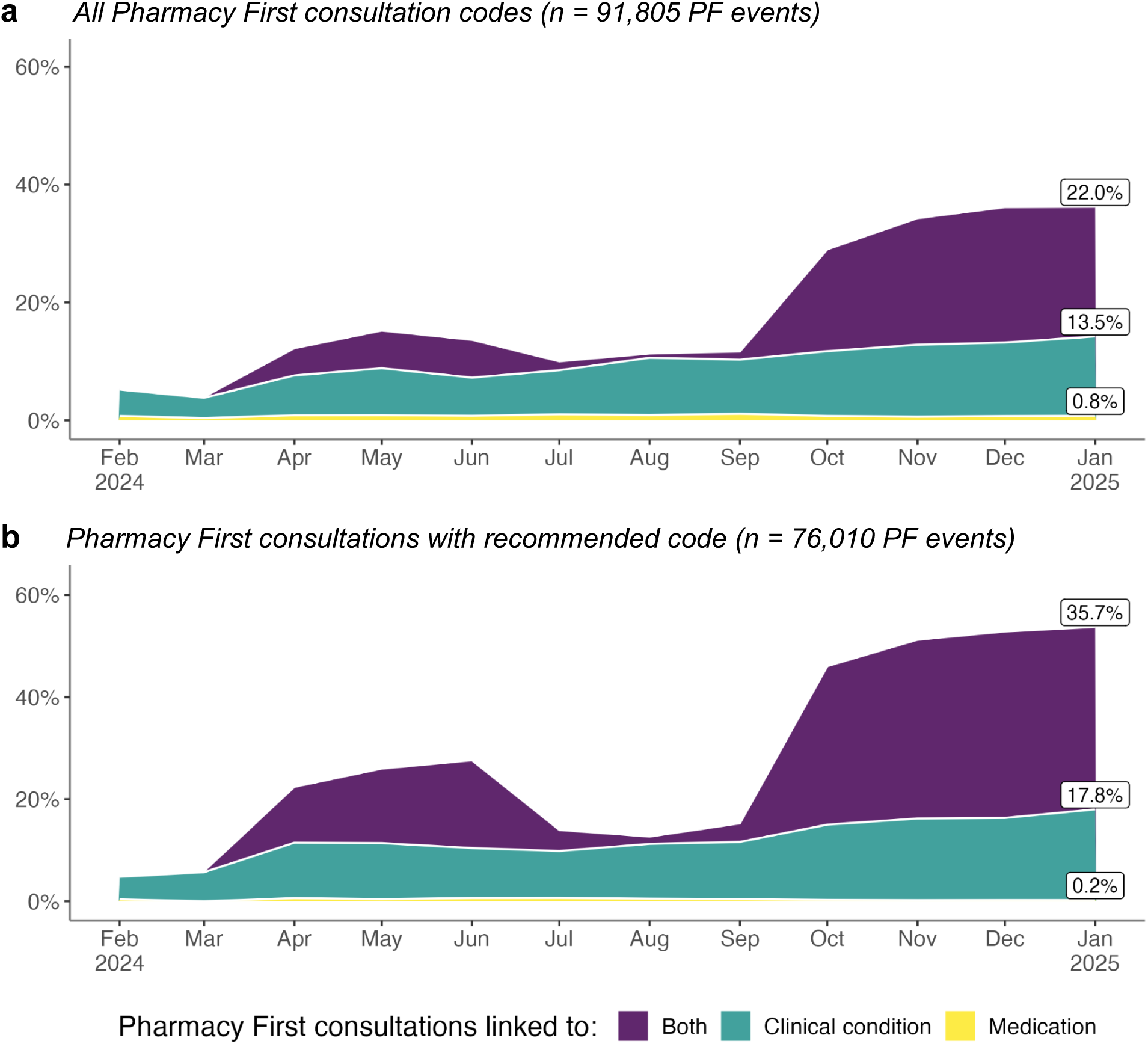
Monthly breakdown of all Pharmacy First consultation codes with either a recorded Pharmacy First medication, clinical condition, or both in OpenSAFELY-TPP, for: **(a)** all consultations with any Pharmacy First consultation code; **(b)** only consultations with the recommended Pharmacy First code - Community Pharmacist Consultation Service for minor illness (1577041000000109).

### Clinical conditions recorded in Pharmacy First consultations

Among consultations with recorded clinical conditions, acute pharyngitis (28.9%) and uncomplicated UTI (28%) were most frequently recorded (Table 2), followed by acute otitis media (16.2%), sinusitis (13.6%), infected insect bites (7.5%), impetigo (2.9%) and herpes zoster (2.8%). All conditions were more commonly recorded among female patients, notably 55 male cases were recorded under the female UTI pathway. These findings closely matched NHS BSA data, with differences ranging from 0.1% (herpes zoster), to 4.5% (acute pharyngitis), see supplementary Table A3.

**Table 2.** Patients with recorded Pharmacy First clinical conditions in OpenSAFELY-TPP during the first 12 months following the services’ launch date on 31 January 2024, grouped by sex.

| Clinical Condition (Inclusion Criteria*) | Sex |  |  |  | Overall | % |
| --- | --- | --- | --- | --- | --- | --- |
|  | Female | % | Male | % |  |  |
| Acute otitis media (1 to 17 years) | 7,365 | 54.8% | 6,075 | 45.2% | 13,440 | 16.2% |
| Acute pharyngitis (5 years and over) | 15,030 | 62.9% | 8,850 | 37.0% | 23,895 | 28.9% |
| Acute sinusitis (12 years and over) | 7,760 | 68.7% | 3,470 | 30.7% | 11,295 | 13.6% |
| Herpes zoster (18 years and over) | 1,470 | 62.8% | 860 | 36.8% | 2,340 | 2.8% |
| Impetigo (1 year and over) | 1,350 | 55.8% | 1,070 | 44.2% | 2,420 | 2.9% |
| Infected insect bites (1 year and over) | 3,955 | 63.5% | 2,270 | 36.5% | 6,225 | 7.5% |
| Uncomplicated UTI (Women 16-64 years) | 23,140 | 99.8% | 55 | 0.2% | 23,195 | 28.0% |
| **Total** | **59,865** | **72.6%** | **22,545** | **27.4%** | **82,410** | **100%** |

Notes. PF = Pharmacy First. UTI = Urinary Tract Infections. \* To capture the entire service activity, sex and age inclusion criteria as specified by NHS England were not applied but are presented for reference. Counts represent distinct patients with $\geq 1$ recorded event for each clinical condition during the study period; a patient may appear in more than one condition category if they had multiple conditions recorded.

**Table 3.**
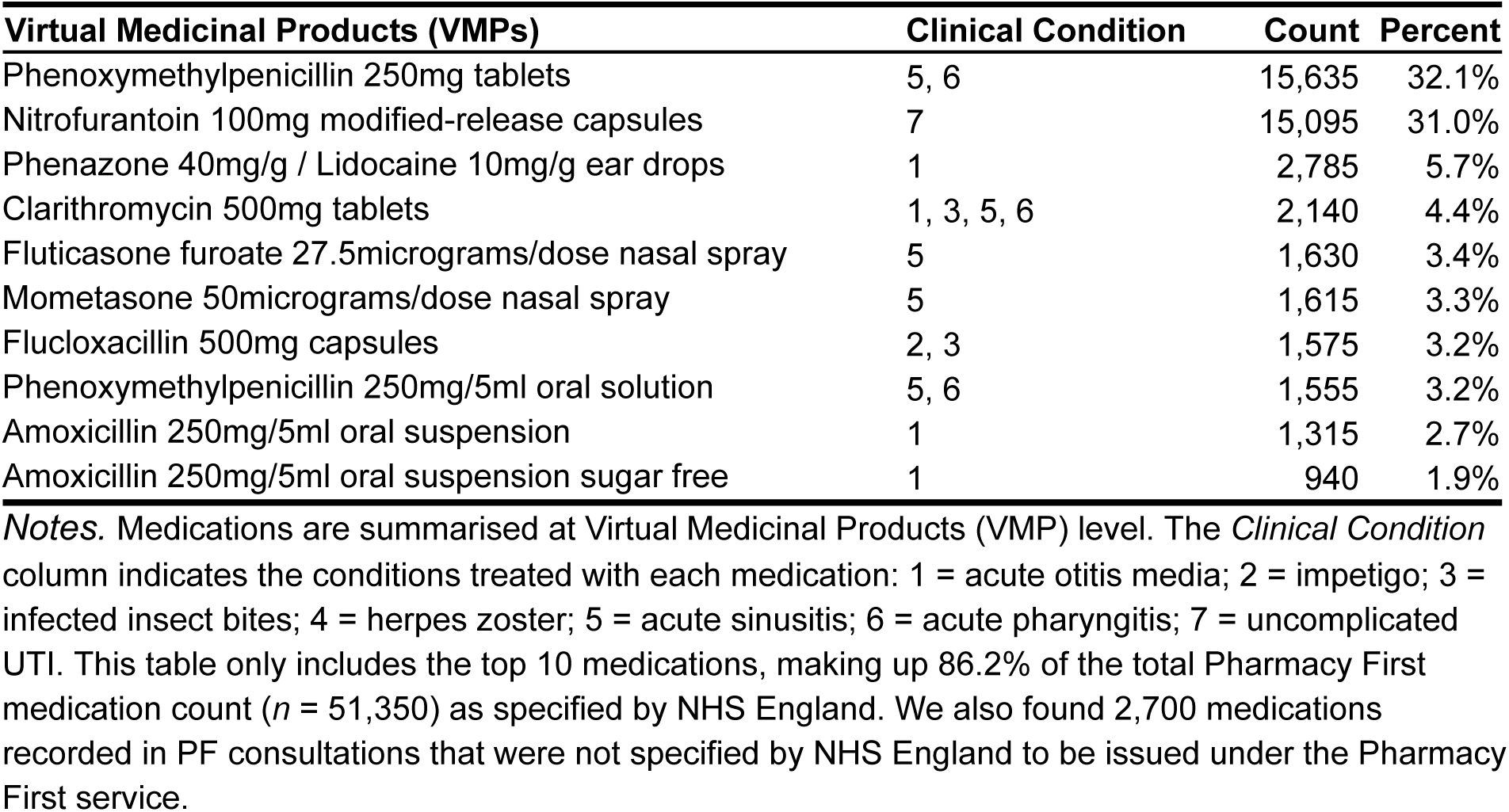
Top ten medication counts and percentage recorded in Pharmacy First consultations during the first 12 months following the services’ launch date on 31 January 2024.

### Medications recorded in Pharmacy First consultations

Among consultations with a recorded medication, the most frequently issued were phenoxymethylpenicillin 250mg tablets (32.1%) and nitrofurantoin 100mg modified-release capsules (31%). Other medications each made up <6% of the total. The top 10 accounted for 86.2% of all medications recorded in Pharmacy First consultations.

## Discussion

### Summary

Pharmacy First was introduced on 31st Jan 2024 following the COVID-19 pandemic. Among 26,142,380 registered patients in OpenSAFELY-TPP, we observed 402,165 Pharmacy first consultations in GP records in its first year, from around 25,000 per month to 56,495 by January 2025. This represented 23.7% of total Pharmacy First consultations reimbursed.

Where present, the most frequently recorded clinical conditions were acute pharyngitis and uncomplicated UTI. By January 2025, 36.3% of Pharmacy First consultations had a recorded clinical condition, medication, or both (53.7% of those with the recommended Pharmacy First code).

### Findings in context

This study provides the first large-scale assessment of recording of Pharmacy First activity in GP records. While published figures report the total monthly Pharmacy First service use with condition/medication breakdowns, the OpenSAFELY platform allows more detailed studies on the patient populations using these services and the potential impacts on patient flows. Here we establish general data quality and patterns of Pharmacy First service use as transferred to practices.

Consultation counts in NHS BSA data were higher, as expected given its complete coverage of reimbursed consultations compared with OpenSAFELY-TPP’s 40% population coverage, but further discrepancies are also present. Pharmacy First consultations are almost all attributed to an identifiable patient (98.94%, February-October 2024) (15) so transfer to their GP records is theoretically possible. Pharmacies may send consultation information by post/email if electronic systems fail, which may not be coded by the receiving practice.

Discrepancies may therefore partly reflect known issues with the *GP Connect: Update Record* functionality (16) and variation in subsequent coding.

The increase in Pharmacy First consultations in OpenSAFELY-TPP between September 2024 and January 2025 likely reflects resolution of technical issues after August 2024 (17), alongside growing awareness and uptake of the Pharmacy First service. Some duplication of records within GP systems is also possible, as we observed fewer unique patients per consultation in OpenSAFELY-TPP (0.847; 340,710/402,165) compared with data from pharmacies (0.963; 2.25 million/2.35 million, April 2024-March 2025) (15).

By January 2025, 36.3% of Pharmacy First consultations in OpenSAFELY-TPP included clinical information; this proportion was higher among consultations recorded using the recommended Pharmacy First code (53.7%), indicating lower levels of recorded clinical information among consultations recorded using non-recommended Pharmacy First codes. Recording a suspected/confirmed clinical condition in a Pharmacy First pathway is not mandatory, and may therefore be absent in GP records (6). However, national data indicate that a medication was issued for around three quarters of all Pharmacy First consultations (18), and by implication, a condition should be confirmed in these cases. The apparent incompleteness of information transferred to GP records may limit further analysis, but information available in a clinical setting may be different.

Pharmacy First service users differed demographically from the overall OpenSAFELY-TPP population. Male patients were underrepresented (32.7% of all Pharmacy First consultations, and 38.1% among those with a recorded clinical condition excluding female urinary tract infection vs 50.2% in OpenSAFELY-TPP). This is consistent with national Pharmacy First data, which reports 36% male consultations (April 2024 - March 2025) (15), and with evidence that men are less likely to consult for common conditions (19). Younger age groups (0-39 years) accounted for the majority of consultations (59.3%). Patients from Yorkshire and the Humber were over-represented (23.7% vs 14.5% in OpenSAFELY-TPP). Patients from more deprived areas were also over-represented, and White British ethnicity also slightly higher (71.3% vs 67.1%). Patterns in deprivation and ethnicity may reflect the greater availability of community pharmacies in deprived areas (20), which could help mitigate aspects of the inverse care law observed in general practice (21,22). However, variation will also reflect underlying clinical condition incidence, healthcare access patterns, and recording practices.

### Strengths and limitations

The main strength of this study is the use of OpenSAFELY-TPP, a secure, population-scale platform covering 26 million patients across 2,540 general practices. This dataset is largely representative of the national population with regards to IMD, age, sex, and ethnicity, making the findings generalisable to England (23). Additionally, this study is the first to evaluate the recording of Pharmacy First consultations in GP records at scale, providing insight into service uptake, data quality, and integration into primary care systems.

Limitations include the inclusive definition of a Pharmacy First consultation used in this study, which incorporates multiple SNOMED CT codes, and may capture some consultations outside the official service specification. London’s underrepresentation in OpenSAFELY-TPP may reduce applicability to urban populations. While OpenSAFELY has been used to deliver analysis across the full English GP population (24,25), access to OpenSAFELY-EMIS (covering the remaining GPs) is currently paused, whilst NHS England concludes formal commercial and data processing agreements with EMIS (26).

### Policy implications

Presence of consultations and clinical details improved over the study period, likely reflecting technical refinements and growing familiarity with the system. Transfer of all Pharmacy First consultations was unlikely during the first year, given the real-world challenges of rapidly implementing national infrastructure. Future services should be supported by timely publication of technical documentation, including SNOMED CT codes to support transparency and independent evaluations. The temporary suspension of *GP Connect: Update Record* functionality during industrial action highlights the need for resilient and clinical safety-assured digital systems. Improved digital integration between community pharmacy and general practice will enhance service monitoring and evaluation, and support the integration of Pharmacy First within routine general practice workflows.

To support ongoing evaluation of equitable access, we developed a public dashboard (https://reports.opensafely.org/) presenting stratified, near real-time data on Pharmacy First consultations. This tool enables policymakers and clinicians to monitor detailed uptake and patterns of Pharmacy First activity over time, supporting oversight of the national roll-out. In October 2025, a large increase in Pharmacy First activity coincided with *GP Connect* becoming a contractual obligation for GPs (27) and addition of some gateway (reimbursement) criteria (17,27).

### Future research

This analysis forms the foundation for a broader mixed methods programme exploring the wider impact of the Pharmacy First service on primary care service use and prescribing (11). This will explore whether Pharmacy First contributes to improving primary healthcare access, the quality of antimicrobial use, and potential scope for refinements to the service, providing robust evidence to inform better implementation and for policymakers to determine the future role of community pharmacy in England.

Further investigation is needed to understand variation in the recording of Pharmacy First consultations, including the use of non-recommended codes, and whether this reflects technical constraints, data entry practices, or limited uptake of *GP Connect: Update Record*. Validation work should also test alignment with pathway inclusion criteria, such as male patients recorded under the UTI pathway or medications outside the service specification.

Qualitative research with pharmacists, GPs, and patients is planned to explore these findings. OpenSAFELY’s tools and infrastructure are openly available, enabling further studies of Pharmacy First and near real-time monitoring of the service’s rollout.

## Conclusion

This study provides the first large-scale analysis of Pharmacy First consultation recording in general practice records. Recorded uptake increased over time, particularly among younger and more deprived populations. Although detailed information was expected to transfer reliably into GP records, the total count of Pharmacy First consultations fell short of the expected total, and some lacked coded clinical information. Strengthening technical integration and ensuring consistent recording across systems will be essential to support robust evaluations of the service and its impact. We invite others to expand on our initial work using OpenSAFELY.

## Administrative

### Funding

The OpenSAFELY platform is principally funded by grants from:

NHS England [2023-2025]; The Wellcome Trust (222097/Z/20/Z) [2020-2024]; MRC (MR/V015737/1) [2020-2021].

Additional contributions to OpenSAFELY have been funded by grants from:

MRC via the National Core Study programme, Longitudinal Health and Wellbeing strand (MC_PC_20030, MC_PC_20059) [2020-2022] and the Data and Connectivity strand (MC_PC_20058) [2021-2022]; NIHR and MRC via the CONVALESCENCE programme (COV-LT-0009, MC_PC_20051) [2021-2024]; NHS England via the Primary Care Medicines Analytics Unit [2021-2024]. This project was funded by a grant from the National Institute for Health and Care Research [NIHR160217].

Diane Ashiru-Oredope is funded by NIHR Senior Clinical and Practitioner Award (NIHR304553).

The views expressed are those of the authors and not necessarily those of the NIHR, NHS England, UK Health Security Agency (UKHSA), the Department of Health and Social Care, or other funders. Funders had no role in the study design, collection, analysis, and interpretation of data; in the writing of the report; and in the decision to submit the article for publication.

### Information governance and ethical approval

NHS England is the data controller of the NHS England OpenSAFELY COVID-19 Service; TPP is the data processor; all study authors using OpenSAFELY have the approval of NHS England.^[3]^ This implementation of OpenSAFELY is hosted within the TPP environment which is accredited to the ISO 27001 information security standard and is NHS IG Toolkit compliant;^[4]^

Patient data has been pseudonymised for analysis and linkage using industry standard cryptographic hashing techniques; all pseudonymised datasets transmitted for linkage onto OpenSAFELY are encrypted; access to the NHS England OpenSAFELY COVID-19 service is via a virtual private network (VPN) connection; the researchers hold contracts with NHS England and only access the platform to initiate database queries and statistical models; all database activity is logged; only aggregate statistical outputs leave the platform environment following best practice for anonymisation of results such as statistical disclosure control for low cell counts.^[5]^

The service adheres to the obligations of the UK General Data Protection Regulation (UK GDPR) and the Data Protection Act 2018. The service previously operated under notices initially issued in February 2020 by the the Secretary of State under Regulation 3(4) of the Health Service (Control of Patient Information) Regulations 2002 (COPI Regulations), which required organisations to process confidential patient information for COVID-19 purposes; this set aside the requirement for patient consent.^[6]^ As of 1 July 2023, the Secretary of State has requested that NHS England continue to operate the Service under the COVID-19 Directions 2020.^[7]^ In some cases of data sharing, the common law duty of confidence is met using, for example, patient consent or support from the Health Research Authority Confidentiality Advisory Group.^[8]^

Taken together, these provide the legal bases to link patient datasets using the service. GP practices, which provide access to the primary care data, are required to share relevant health information to support the public health response to the pandemic, and have been informed of how the service operates.

This study was supported by Anne Joshua (Deputy Director of Pharmacy Commissioning & Head of Pharmacy Integration, NHS England) as senior sponsor, and approved by the Health Research Authority (REC reference 20/LO/0651). (NHS England service evaluations/audits are currently not required to have Ethics approval.)

1. NHS Digital. The NHS England OpenSAFELY COVID-19 service - privacy notice [Internet]. 2023 [cited 2023 Jul 4]. Available from: https://digital.nhs.uk/coronavirus/coronavirus-covid-19-response-information-governance-hub/the-nhs-england-opensafely-covid-19-service-privacy-notice
2. NHS Digital. Secretary of State for Health and Social Care - UK Government. COVID-19 Public Health Directions 2020: notification to NHS Digital [Internet]. 2023 [cited 2023 Jul 4]. Available from: https://web.archive.org/web/20250414105355/https://digital.nhs.uk/about-nhs-digital/corporate-information-and-documents/directions-and-data-provision-notices/data-provision-notices-dpns/opensafely-covid-19-service-data-provision-notice
3. NHS Digital. ISB1523: Anonymisation Standard for Publishing Health and Social Care Data [Internet]. 2023 [cited 2023 Jul 4]. Available from: https://web.archive.org/web/20250430135246/standards.nhs.uk/published-standards/anonymisation-standard-for-publishing-health-and-social-care-data
4. NHS Digital. Data Security and Protection Toolkit [Internet]. 2023 [cited 2023 Jul 4]. Available from: https://web.archive.org/web/20250405100349/https://digital.nhs.uk/services/data-security-and-protection-toolkit/data-security-and-protection-toolkit
5. NHS Digital. Coronavirus (COVID-19): notice under regulation 3(4) of the Health Service (Control of Patient Information) Regulations 2002 – general [Internet]. 2022 [cited 2023 Jul 5]. Available from: https://www.gov.uk/government/publications/coronavirus-covid-19-notification-of-data-controllers-to-share-information/coronavirus-covid-19-notice-under-regulation-34-of-the-health-service-control-of-patient-information-regulations-2002-general--2
6. Health Research Authority. Confidentiality Advisory Group [Internet]. 2023 [cited 2023 Jul 4]. Available from: https://web.archive.org/web/20250408154826/https://www.hra.nhs.uk/about-us/committees-and-services/confidentiality-advisory-group/

## Data Availability

All data were linked, stored, and analysed securely within the OpenSAFELY platform (https://opensafely.org). Data include pseudonymised information such as coded diagnoses, medications, and consultation details. No free-text data were included.
Detailed pseudonymised patient data are potentially re-identifiable and cannot be shared. All analysis code is openly available for review and reuse under the MIT open licence at https://github.com/opensafely/pharmacy-first.
This study also used publicly available data from the NHS Business Services Authority (NHS BSA) on dispensing contractors, available under the Open Government Licence at: https://www.nhsbsa.nhs.uk/prescription-data/dispensing-data/dispensing-contractors-data
.

https://github.com/opensafely/pharmacy-first

## Acknowledgements

We are very grateful for all the support received from the TPP Technical Operations team throughout this work, and for generous assistance from the information governance and database teams at NHS England and the NHS England Transformation Directorate.

