## Supplementary Materials for "Recording of Pharmacy First consultations in general practice records in England: an observational study of the service’s first year using OpenSAFELY"

### Online Supplemental Materials

**Table A1**. Codelists used to identify Pharmacy First consultations, clinical events, and medications.

| Category | Codelist |
|---|---|
| **PF Consultations** | opensafely/pharmacy-first-consultation |
| **PF Clinical Conditions** | user/chriswood/pharmacy-first-clinical-pathway-conditions |
| **PF Medications** | |
| Acute Otitis Media | opensafely/pharmacy-first-acute-otitis-media-treatment-full-dmd-codelist |
| Impetigo | opensafely/pharmacy-first-impetigo-treatment-full-dmd-codelist |
| Infected Insect Bites | opensafely/pharmacy-first-infected-insect-bites-treatment-full-dmd-codelist |
| Herpes Zoster | codelist/opensafely/pharmacy-first-shingles-treatment-full-dmd-codelist |
| Acute Sinusitis | codelist/opensafely/pharmacy-first-sinusitis-treatment-full-dmd-codelist |
| Acute Pharyngitis | opensafely/pharmacy-first-sore-throat-treatment-full-dmd-codelist |
| UTI | opensafely/pharmacy-first-urinary-tract-infection-treatment-full-dmd-codelist |

*Notes.* PF = Pharmacy First; UTI = Urinary Tract Infections.

**Table A2.** Count of clinical conditions linked to Pharmacy First consultations in OpenSAFELY-TPP grouped by IMD during the first 12 months following the services' launch.

| **Clinical Condition (Inclusion Criteria*)** | **Index of Multiple Deprivation (IMD)** | | | | |
|---|---|---|---|---|---|
| | 1 (most deprived) | 2 | 3 | 4 | 5 (least deprived) |
| Acute otitis media (1 to 17 years) | 3,305 | 2,305 | 2,315 | 2,360 | 2,055 |
| Acute pharyngitis (5 years and over) | 6,685 | 5,070 | 4,145 | 3,710 | 3,040 |
| Acute sinusitis (12 years and over) | 2,335 | 1,970 | 2,215 | 2,240 | 1,940 |
| Herpes zoster (18 years and over) | 410 | 380 | 460 | 505 | 450 |
| Impetigo (1 year and over) | 630 | 445 | 415 | 435 | 360 |
| Infected insect bites (1 year and over) | 1,335 | 1,150 | 1,255 | 1,215 | 1,000 |
| Uncomplicated UTI (Women 16-64 years) | 5,295 | 4,300 | 4,390 | 4,350 | 3,680 |

*Notes.* UTI = Urinary Tract Infections.* To capture the entire service activity, inclusion criteria as specified by NHS England were not applied but are presented for reference.

**Table A3.** Comparison of clinical condition recorded in Pharmacy First consultations in OpenSAFELY-TPP and NHS BSA during the first 12 months following the services' launch.

| **Clinical Condition (Inclusion Criteria*)** | **OpenSAFELY-TPP** | | **NHS BSA** | |
|---|---|---|---|---|
| | Count** | Percent | Count** | Percent |
| Acute otitis media (1 to 17 years) | 13,440 | 16.2% | 260,590 | 11.8% |
| Acute pharyngitis (5 years and over) | 23,895 | 28.9% | 736,647 | 33.4% |
| Acute sinusitis (12 years and over) | 11,295 | 13.6% | 248,702 | 11.3% |
| Herpes zoster (18 years and over) | 2,340 | 2.8% | 58,361 | 2.7% |
| Impetigo (1 year and over) | 2,420 | 2.9% | 736,647 | 4.3% |
| Infected insect bites (1 year and over) | 6,225 | 7.5% | 198,069 | 9.0% |
| Uncomplicated UTI (Women 16-64 years) | 23,195 | 28.0% | 607,646 | 27.5% |

*Notes.* UTI = Urinary tract infections. * To capture the entire service activity, inclusion criteria as specified by NHS England were not applied but are presented for reference.
** OpenSAFELY-TPP counts in this table represent one condition per patient; NHS BSA counts capture all observed conditions, including multiple per patient. For the purpose of this comparison, we assume that this does not significantly impact the proportional representation of each condition.
